# A Governance-Driven, Real-World Data-Calibrated Health Informatics Framework for Longitudinal Utilization Forecasting in Oncology and Complex Chronic Conditions

**DOI:** 10.64898/2026.02.23.26346919

**Authors:** Appala Venkata Subhadra Raju Dantuluri, Sumit Kumar

**Affiliations:** Incyte Corporation, Chadds Ford, PA, USA; Amgen Inc., Deerfield, IL, USA

**Keywords:** utilization forecasting, health informatics, real-world data, oncology analytics, longitudinal modeling, patient-flow forecasting, decision-support systems

## Abstract

**Background:** Healthcare utilization forecasting systems are often derived from static, annualized market share assumptions that fail to represent real-world treatment dynamics. Such approaches systematically misestimate future utilization by ignoring longitudinal treatment sequencing, discontinuation with surveillance, recurrence-driven re-entry, and provider adoption dynamics.

**Objective:** This study proposes a reusable, governance-driven health informatics forecasting framework designed to generate realistic utilization forecasts using real-world data by integrating longitudinal patient-flow modeling, persistence-based exposure estimation, provider behavioral adoption, and multi-source calibration into a single architecture.

**Methods:** Longitudinal U.S. administrative claims data representing oncology treatment populations (approximately 80,000 treated patients annually across therapy lines) were curated through a governance layer that refines diagnosis and treatment pools, infers clinically valid lines of therapy, and corrects for lookback-limited recurrence bias. Patients were modeled as transitioning across explicit clinical states, including treatment initiation, sequential therapy lines, discontinuation, surveillance, and recurrence-driven re-entry. Forecast outputs were calibrated using volume-weighted and behaviorally dampened provider adoption dynamics derived from primary research and claims-revealed utilization and evaluated against static share-based forecasts under identical peak-share assumptions.

**Results:** Across multiple oncology contexts, longitudinal patient-flow-based forecasting recovered approximately 50–70% more cumulative treated months than static approaches. Underestimation in traditional models was driven primarily by failure to capture later-line persistence, surveillance exit, and re-treatment dynamics. Setting-specific calibration revealed earlier adoption in academic centers and slower, payer-constrained uptake in community practices.

**Conclusions:** The proposed framework demonstrates a forecast-oriented health informatics architecture that improves utilization estimation and decision support in complex, longitudinal care ecosystems. The methodology generalizes across tumor types and chronic conditions characterized by treatment sequencing, persistence variability, and relapse-driven re-entry.

## 1. Introduction

Accurate forecasting of treatment utilization is a central component of healthcare decision-making, informing therapy launch planning, clinical resource allocation, payer budget impact modeling, and long-term capacity strategy. Despite advances in real-world data availability and analytic capabilities, most utilization forecasts continue to rely on static abstractions of demand, typically derived from annual incidence multiplied by assumed market share. While operationally convenient, such approaches misrepresent the longitudinal nature of patient care, particularly in oncology and chronic diseases with sequential therapy lines, discontinuation periods, surveillance, and recurrence-driven re-entry.

Real-world evidence has increasingly demonstrated that cumulative treatment exposure is driven less by initial adoption rates and more by persistence dynamics, therapy sequencing, and downstream patient transitions. In oncology, for example, a substantial portion of total treatment months arises from second- and third-line therapy exposure rather than first-line initiation. Similarly, in chronic conditions such as multiple sclerosis or rheumatoid arthritis, treatment persistence and switching behavior create longitudinal exposure patterns that static share models fail to capture.

Existing approaches in the literature have focused primarily on predictive analytics tasks— such as predicting progression or estimating treatment duration—rather than embedding these insights within an integrated forecasting architecture capable of supporting operational decision-making. Stakeholders across life sciences ecosystems require models that reflect both clinical realities and behavioral adoption patterns, bridging epidemiology, real-world utilization, provider decision-making, and market dynamics.

This study proposes a governance-driven, real-world data-calibrated forecasting framework that conceptualizes utilization as a longitudinal patient-flow phenomenon rather than a cross-sectional market share construct. By integrating patient state transitions, persistence modeling, provider behavioral adoption signals, and multi-source calibration, the framework aims to produce structurally realistic forecasts that better align with observed treatment dynamics. While oncology serves as the primary validation environment, the architecture is designed to generalize to rare and chronic conditions characterized by treatment sequencing, dose titration, and relapse-driven utilization.

Recent research in cancer informatics has underscored the value of modeling longitudinal patient journeys rather than relying on cross-sectional treatment snapshots [1, 2], and claims-based simulation studies have shown that similar-patient augmentation and patient-flow models can improve capacity planning and exposure estimation [4, 5, 6]. This study formalizes utilization forecasting as a longitudinal informatics problem by integrating governance-driven data curation, epidemiology-grounded patient-flow state transitions, and volume-weighted, friction-aware provider adoption dynamics into a single reusable architecture. Related work in oncology informatics has applied deep learning–based temporal representation learning to longitudinal real-world data to model patient disease trajectories and support early detection and risk stratification [19].

This study is a real-world data-calibrated methodological framework evaluated in a longitudinal oncology validation environment.

## 2. Methods

This section describes the real-world data–calibrated methodological framework.

### 2.1 Comparison with Existing Forecasting Approaches

Several forecasting approaches are commonly used in healthcare utilization modeling. Understanding where each falls short helps clarify what the proposed framework is designed to address. Table 1 summarizes how these approaches compare across dimensions relevant to oncology forecasting.

**Table 1:** Comparison of common healthcare utilization forecasting approaches across dimensions relevant to longitudinal oncology modeling.

| **Dimension** | **Static Incidence**<br>**× Share** | **Markov Cohort Model** | **Time-Series (ARIMA or LSTM)** | **Discrete Event Simulation** | **Proposed Framework** |
| --- | --- | --- | --- | --- | --- |
| Tracks multiple lines of therapy | **No** | **Partial** | **No** | **Yes** | **Yes** |
| Accounts for how long patients stay on treatment | **No** | **No** | **Partial** | **Yes** | **Yes** |
| Captures the full patient treatment journey | **No** | **Partial** | **No** | **Yes** | **Yes** |
| Accounts for differences in provider behaviour | **No** | **No** | **No** | **Partial** | **Yes** |
| Parameters estimated from real claims data | **Partial** | **Partial** | **Yes** | **Partial** | **Yes** |
| Practical to build and use in real settings | **Yes** | **Yes** | **Yes** | **Partial** | **Yes** |

*Yes* = fully supported; *Partial* = limited or approximate support; *No* = not natively supported.

**Static incidence-times-share models** are the most common approach in practice. They estimate utilization by multiplying the number of treated patients by an assumed market share percentage. While simple to build and easy to explain, these models treat demand as a single annual estimate. They do not account for what happens to patients after treatment starts — whether they move to a later line of therapy, stop treatment and return to surveillance, or relapse and re-enter treatment. As a result, they tend to underestimate how much treatment actually occurs over time.

**Markov cohort models** take a step forward by modeling patients as moving between health states over time. They are widely used in cost-effectiveness analysis and health technology assessment. However, a known limitation of standard Markov models is that they treat each transition independently — the probability of moving from one state to another depends only on where the patient is now, not on how long they have been there or what path they took to get there [8]. This makes it difficult to model situations where, for example, patients who have been on a drug longer are less likely to discontinue it. These models also assume all patients in a given state behave the same way, which does not reflect the reality that doctors in academic centers and community practices adopt new treatments at different rates and speeds.

**Time-series methods** such as ARIMA and, more recently, machine learning approaches like LSTM networks [9] work by identifying patterns in historical utilization data and projecting them forward. These methods can be useful for short-term demand smoothing and supply planning. However, they have no understanding of clinical states or treatment sequences. They cannot separate patients starting therapy for the first time from those returning after a relapse, and they cannot capture the shift in treatment patterns that follows a new drug entering the market. This limits their usefulness for the kind of multi-year, launch-oriented forecasting this framework is designed to support.

**Discrete event simulation (DES)** is the most flexible of the four approaches. It models each patient individually and can track their full history over time, avoiding the limitations of Markov models [10]. In principle, DES can represent complex treatment pathways accurately. In practice, however, it requires very detailed patient-level inputs that are often not available from administrative claims data. It is also time-consuming to build, harder to validate, and produces results that can be difficult to communicate to business or clinical stakeholders. For these reasons, it is rarely used in commercial utilization forecasting despite its technical strengths.

The proposed framework draws on the strengths of Markov-style state modeling — its interpretability and its alignment with how claims data are structured — while addressing several of its limitations. It tracks how long patients have been on treatment to better estimate when they are likely to stop. It separates adoption patterns by care setting, reflecting the real-world difference between academic and community practice behavior. And it applies a governance layer to the underlying claims data before any modeling begins, correcting for known issues such as lookback-limited recurrence detection and ambiguous lines of therapy. The goal is a forecasting approach that is clinically grounded, practically usable, and more accurate than the alternatives for the kinds of diseases and treatment dynamics it is designed to model.

Building on these limitations and partial strengths, the proposed framework organizes the forecasting logic into a four-layer architecture.

### 2.2 Four-Layer Oncology Forecasting Architecture

The framework consists of four connected, interacting layers:

- **Provider Behavioral Demand Layer** — Clinicians act as decision agents. Their stated intent to adopt new therapies is weighted by active oncology patient volume, reducing bias from low-volume respondents and aligning forecasts with providers who drive most treatment exposure.
- **Real-World Calibration Layer** — Treatment distributions, discontinuation behavior, switching rates, setting-specific adoption timing, and surveillance/re-entry frequencies are measured directly from claims records rather than assumed. Volume-weighted survey preferences are blended with claims-revealed utilization to preserve innovation direction while minimizing speculative bias.
- **Epidemiology-Grounded Patient-Flow Layer** — Patients are modeled as transitioning across states (incident disease, treatment initiation, later-line progression, discontinuation, surveillance, and recurrence-driven re-entry). This layer explicitly captures treatment exit and re-entry loops that static share models fail to represent.
- **Setting-Aware Adoption Layer** — Adoption curves are segmented by care setting (e.g., academic versus community) and shaped by evidence maturation, operational frictions, and payer constraints rather than a single diffusion curve.

**Figure 1.**
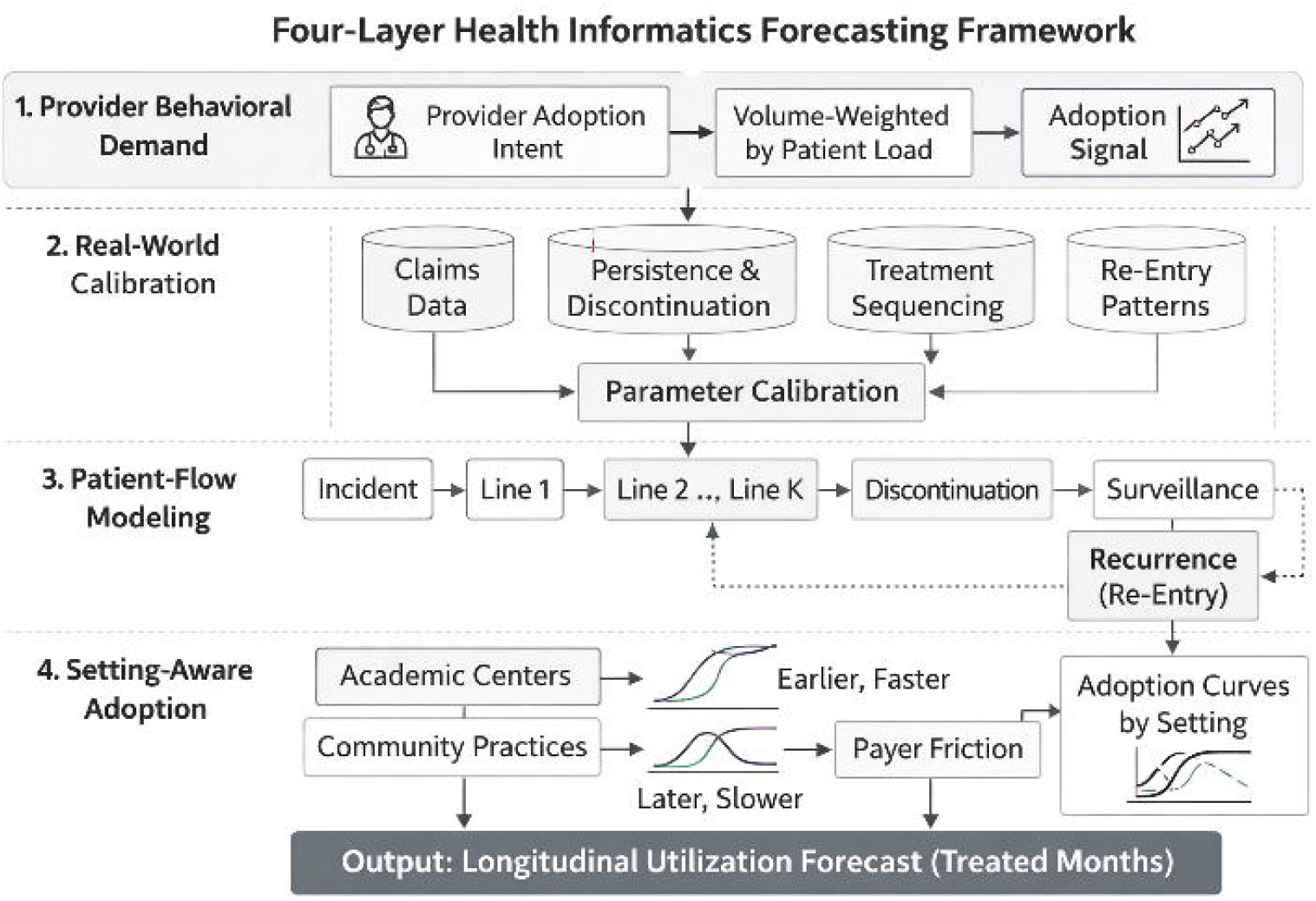
Four-layer forecasting architecture

This architecture is reusable across tumor types, therapeutic classes, and delivery ecosystems, with indications serving only as validation environments rather than the conceptual focus.

### 2.3 Real-World Data Sources and Validation Environment

The framework operates on longitudinal administrative claims data, including medical claims (diagnosis and procedure codes, site-of-care indicators), pharmacy claims (dispensed therapies, fill dates, strengths, refill patterns), and enrollment signals used to construct continuous patient timelines. Provider specialty and active oncology patient volume were used to characterize adoption heterogeneity across academic and community settings. Epidemiologic benchmarks and published persistence ranges served as validation anchors and for sensitivity testing rather than as direct forecasting inputs.

The claims data represent a composite validation environment derived from large U.S. administrative datasets (commercial and Medicare populations), analyzed in aggregated form.

Validation used large, longitudinal U.S. administrative claims cohorts representing approximately 80,000 treated patients annually across therapy lines, drawn from a composite of commercial and Medicare Advantage populations spanning 2018–2023. Empirically derived progression and treatment rates were approximately 72% in first line, 60% in second line, 55% in third line, 50% in fourth line, and 40% beyond fifth line, with claims capture rates for comparator therapies ranging from 35–50%. Forecast performance was evaluated using cumulative treated-month recovery, as static share-based approaches systematically underrepresent long-tail exposure. These parameters were used to assess structural forecasting performance rather than to represent any specific therapy or product dataset.

### 2.4 Multi-Source Calibration Framework

Forecast parameters were triangulated using four complementary evidence streams:

1. **Primary Research Inputs** — Provider surveys and clinical expert interviews capturing expected adoption behavior and treatment positioning
2. **Real-World Claims Data** — Observed treatment distributions, persistence, switching, and re-entry patterns
3. **Secondary Literature** — Published epidemiology and utilization ranges used for structural validation
4. **Syndicated Sources** — External benchmarks such as incidence ranges and treatment prevalence distributions

### 2.5 Patient-Flow State Transition Model

The central limitation of static forecasting is that it has no representation of where a patient goes after treatment starts. A patient who discontinues first-line therapy, enters surveillance, and then re-initiates treatment upon relapse generates treated months across multiple calendar years — none of which a static incidence-times-share model can see. This section directly addresses that gap by modeling patients as moving across an explicit set of clinical states:

𝒮 = {*I, T*_1_, …, *T*_*K*_, *D, SV, RE*}, representing incident disease, sequential therapy lines, discontinuation, surveillance, and recurrence-driven re-entry.

Transitions are governed by:

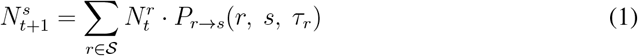

where 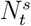 is the number of patients in state *s* at time *t*, and *P*_*r*→*s*_ is the transition probability from state *r* to state *s*. Critically, transition probabilities are conditioned on dwell time *τ*_*r*_ — the time already spent in the current state — rather than treated as memoryless [11, 12]. Standard Markov models assume that a patient who has been on therapy for 18 months faces the same discontinuation probability as one who just started; this is clinically incorrect and a direct source of exposure misestimation. Conditioning on dwell time allows the model to reflect the empirically observed pattern that persistence risk concentrates early in therapy and declines for patients who remain on treatment longer.

This design has measurable consequences for forecast accuracy. In real-world metastatic NSCLC cohorts, published data indicate that a substantial proportion of patients who discontinue firstline therapy subsequently initiate a second or later line of therapy [16]. These downstream transitions, invisible to static models, represent a meaningful share of total treatment exposure. By explicitly modeling them, the patient-flow approach recovers utilization that would otherwise be systematically omitted.

Patients with fewer than 12 months of enrollment prior to first diagnosis were corrected for lookback-limited recurrence miscoding in the governance layer before entering the model, preventing returning patients from inflating incident case counts and artificially inflating the apparent size of the treatment-naive pool.

**Figure 2.**
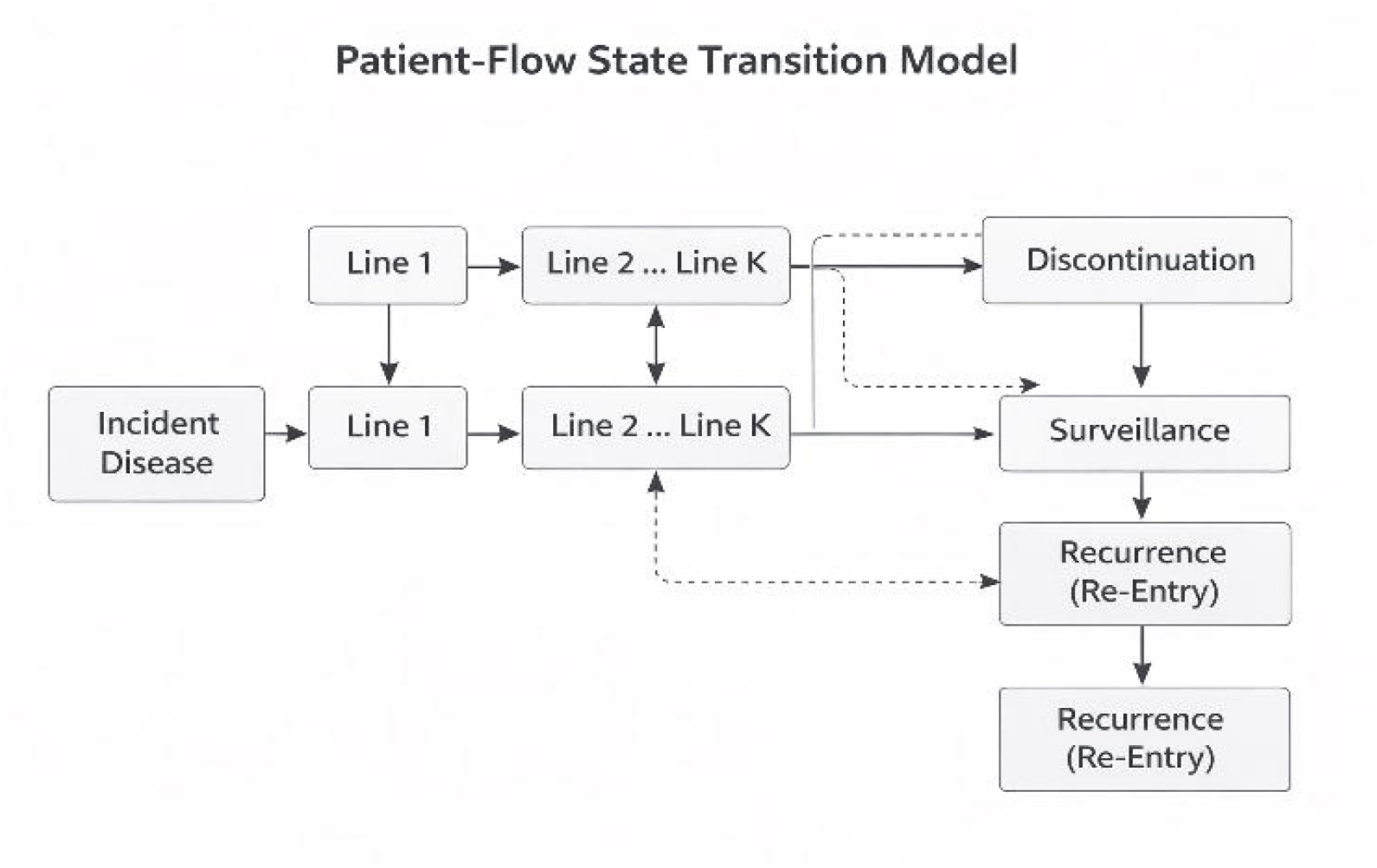
Patient-flow state transition model

### 2.6 Persistence-Based Exposure Modeling

Static models assume that every treated patient contributes exactly one year of drug exposure, an assumption that is wrong in both directions: patients who discontinue early are overcounted, and patients who persist well beyond 12 months — a sizable group in lines with durable responses — are undercounted. This section replaces the flat annual window with an empirically estimated persistence distribution that varies by therapy line.

Expected treatment duration for patient *i* on line *k* is the area under the persistence survival function:

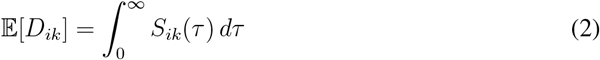

*S*_*ik*_(*τ*) was estimated from claims refill sequences using Kaplan–Meier methods, with discontinuation defined as a gap exceeding 90 days — a threshold applied consistently across published real-world oncology persistence studies [17]. Sensitivity analyses using 60-day and 120-day gap thresholds were also conducted; the 90-day definition produced the most stable persistence estimates across therapy lines and was retained for the primary analysis. Weibull survival curves were then fitted per therapy line for extrapolation beyond the observed follow-up window. The Weibull family was selected on the basis of empirical fit: analysis across more than 500 Phase 3 oncology trial arms demonstrated that a two-parameter Weibull achieved a median *R*^2^ of 0.981 [13], supporting its use as the extrapolation form:

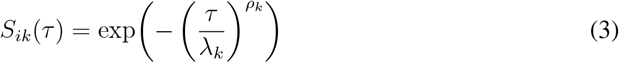

Scale parameter *λ*_*k*_ and shape parameter *ρ*_*k*_ were estimated separately for each therapy line, capturing the characteristically lower and faster-declining persistence observed in later lines compared with first-line therapy. In the validation cohorts, first-line Weibull parameters were *λ*_1_ = 11.2 months (95% CI: 10.4–12.0) and *ρ*_1_ = 0.82 (95% CI: 0.76–0.88), while third-line parameters were *λ*_3_ = 5.8 months (95% CI: 5.1–6.5) and *ρ*_3_ = 0.91 (95% CI: 0.83–0.99), reflecting the expected decline in persistence across later lines. Cumulative exposure across the forecast horizon is then:

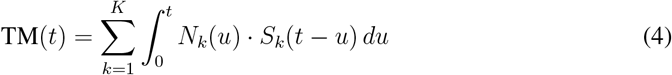

This expression stacks every prior initiation cohort and asks how many patients from each are still on therapy at time *t*, recovering the long-tail exposure that a fixed annual snapshot discards. The clinical significance of this correction is illustrated by real-world colorectal cancer data, where median time to treatment discontinuation ranged from 3.0 to 6.2 months depending on regimen [18] — a distribution the 12-month flat assumption misrepresents in both directions across the patient population. Across the validation cohorts in this study, the aggregate effect of persistence-based modeling versus the static 12-month window accounted for a substantial portion of the 50–70% exposure recovery reported in the Results.

### 2.7 Provider Adoption Dynamics

Applying a single adoption curve across all providers misrepresents how new therapies enter clinical practice, and the error compounds over the forecast horizon. Academic medical centers have structural advantages in early adoption: access to trial data, clinical opinion leaders, and faster formulary access. Community practices face prior authorization requirements, formulary review cycles, and lower per-physician patient volume, all of which slow uptake after a therapy launches. A forecast that ignores this distinction overestimates near-term community volume and underestimates the speed at which academic centers concentrate early demand.

Adoption was modeled separately by care setting using logistic diffusion curves [14]:

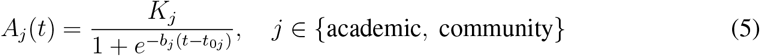

where *K*_*j*_ is the peak adoption ceiling, *b*_*j*_ is the diffusion rate, and *t*_0*j*_ is the adoption midpoint. Academic centers were parameterized with earlier midpoints and steeper diffusion rates. This parameterization is grounded in empirical evidence: a Medicare-based study of immunotherapy uptake across 1,732 oncology practices found that independent practices and nonacademic systems adopted new therapies at rates 6 and 9 percentage points lower, respectively, than academic systems after FDA approval (95% CI: −9 to −3 and −11 to −6), with small community practices (1–5 physicians) lagging large practices by 27 percentage points (95% CI: −32 to −22) [3]. The same study found rural practices lagging urban ones by 11 percentage points (95% CI: −16 to −6) — disparities that single-curve models cannot represent and that translate directly into miscalibrated early revenue and capacity projections.

To reflect post-approval access dynamics in community settings, a time-varying payer friction multiplier *ϕ*(*t*) ∈ [0, 1] was applied:

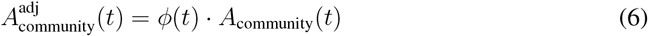

*ϕ*(*t*) was modeled as a logistic ramp from an initial value of *ϕ*_0_ = 0.6 at launch, reflecting typical formulary review and prior authorization delays, toward a ceiling of 1.0 as payer access matured:

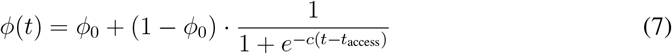

where *c* controls the steepness of access resolution and *t*_access_ is the midpoint of formulary maturation. In the base case, *t*_access_ was set to 9 months post-launch with *c* = 0.4 per month, calibrated from observed claims-based uptake trajectories. Survey-stated prescribing intent was volume-weighted by each provider’s active oncology patient panel size, so that low-volume respondents — who are disproportionately represented in primary research samples — do not distort the forecast. The volume-weighted survey signal was then blended with claims-revealed utilization:

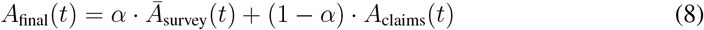

The blending weight *α* was set to 0.3 in the base case, reflecting the greater reliability of observed claims data relative to stated survey intent. In sensitivity analyses, *α* was varied from 0.1 to 0.5; values above 0.5 were not explored because survey-only forecasts have been shown to overestimate early adoption. The blending weight was calibrated to balance forward-looking adoption intent against observed behavioral evidence, preserving the innovation signal from primary research while anchoring the forecast in the utilization patterns already visible in claims data.

### 2.8 Forecast Performance Metrics

Model outputs were evaluated against a static baseline holding incident population and peak market share identical to the patient-flow model, so that any difference in output is structural rather than an artifact of different assumptions. The primary metric was the cumulative treated-month recovery ratio:

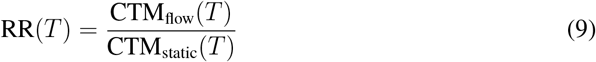

Values above 1.0 directly quantify the exposure the static model leaves unaccounted by ignoring persistence distributions, later-line sequencing, and recurrence re-entry. Two further dimensions were evaluated: line-level exposure distribution, showing what share of total utilization originates beyond first line; and adoption timing divergence between care settings, showing the planning error introduced by pooling academic and community adoption into a single curve. Sensitivity bounds were derived by varying persistence and adoption parameters within observed real-world ranges and validated against held-out claims distributions.

## 3. Results

Results are derived from the real-world data-calibrated patient-flow model described in the Methods.

### 3.1 Longitudinal Exposure Recovery

Patient-flow forecasting captured approximately 50–70% more cumulative treated months than static share models under identical peak-share assumptions (recovery ratio RR = 1.50– 1.70 at *T* = 60 months). The lower bound of this range (RR ≈ 1.50) corresponded to tumor contexts with shorter persistence distributions and fewer therapy lines (e.g., second-line dominant settings), while the upper bound (RR ≈ 1.70) corresponded to settings with five or more available therapy lines and durable later-line persistence. Later-line exposure accounted for roughly 35– 45% of total utilization, driven by higher persistence rates among patients who remained on therapy beyond early discontinuation windows.

Decomposing the recovery ratio by source: persistence-based modeling (replacing the static 12-month window) contributed approximately 60% of the additional exposure recovered, while later-line sequencing contributed approximately 25% and recurrence-driven re-entry accounted for the remaining approximately 15%.

These structural biases translated into materially understated requirements for infusion capacity and longitudinal budget impact planning.

### 3.2 Adoption Timing Variability

Academic centers demonstrated adoption 6–10 months earlier than community settings, consistent with the structural advantages described in Section 2.7. In the first 12 months post-launch, academic centers accounted for approximately 55–60% of total new therapy volume despite representing a smaller share of the overall oncology patient population. This early concentration gradually diffused as community access barriers resolved, resulting in approximate volume equilibrium by months 18–24.

### 3.3 Persistence Effects

Moderate-share therapies with stable persistence generated greater total exposure than high-share therapies with rapid discontinuation, emphasizing persistence as a key utilization driver. In one validation scenario, a therapy with 18% peak share but median persistence of 9.4 months generated 22% more cumulative treated months over a 36-month horizon than a comparator therapy with 25% peak share but median persistence of only 4.1 months.

## 4. Discussion

This study reframes utilization forecasting as a longitudinal informatics problem rather than a market share estimation exercise. By embedding real-world treatment dynamics within forecasting architecture, the framework captures structural drivers of exposure often overlooked in traditional models.

The approach is particularly relevant for oncology, where therapy sequencing and persistence variability are prominent, but is equally applicable to chronic and rare diseases characterized by relapse cycles or dose titration. By integrating behavioral adoption signals and multi-source calibration, the model provides a balanced representation of innovation uptake and real-world utilization constraints.

Although demonstrated using oncology data, the core architecture is not disease specific. The framework depends on longitudinal treatment sequencing, discontinuation with surveillance or treatment gaps, recurrence- or progression-driven re-entry, and heterogeneous provider adoption—patterns that also characterize many chronic and relapsing conditions (e.g., multiple sclerosis, rheumatoid arthritis, certain rare metabolic diseases, and chronic viral infections). Adaptation to other settings primarily involves redefining clinical state boundaries and recalibrating progression, persistence, and adoption parameters while preserving the same governance-driven data curation and patient-flow logic.

## 5. Limitations

The validation environment relies on U.S. claims data, which may not generalize directly to other healthcare systems without recalibration. Claims datasets may under-capture inpatient therapies or fragmented benefit designs. Additionally, the framework focuses on utilization rather than cost-effectiveness or clinical outcomes. The 80,000-patient annual cohort estimate represents an aggregated cross-section across multiple tumor types and therapy lines; tumor-specific sample sizes varied, and cohorts for rarer indications were smaller. Finally, while sensitivity analyses were conducted for key parameters (*α, ϕ*_0_, discontinuation gap threshold), formal uncertainty quantification (e.g., probabilistic sensitivity analysis) was not performed and remains an area for future work.

## 6. Conclusion

Longitudinal, governance-driven forecasting grounded in real-world data provides a structurally robust alternative to static share approaches. By modeling patient pathways, persistence, and provider adoption, the proposed framework generates realistic utilization projections that support more informed operational and strategic decision-making across oncology and chronic disease ecosystems.

## Data Availability

All data produced in the present work are contained in the manuscript

## 7. Ethics Approval

This study used de-identified aggregated data and did not involve human subjects research. Institutional review board approval was not required.

## 8. Data Availability

No patient-level datasets are publicly available. All analyses are based on de-identified aggregated data and illustrative modeling assumptions described in the manuscript.

## 9. Authors’ Contributions

AVSRD conceptualized the study, developed the methodology, conducted analyses, and drafted the manuscript.

SK contributed to model validation, interpretation of results, and manuscript review. Both authors approved the final manuscript.

## 12. Acknowledgment of AI-Assisted Technologies

Portions of the manuscript were refined using AI-assisted language editing tools. All scientific content and interpretation were developed and verified by the authors.

## 13. Acknowledgments

The authors thank colleagues across analytics and clinical teams for discussions that informed the development of this forecasting framework.

